# Joint models reveal genetic architecture of transitions between pubertal stages and their association with BMI in a Latino population

**DOI:** 10.1101/2023.06.29.23292039

**Authors:** Lucas Vicuña, Esteban Barrientos, Valeria Leiva-Yamaguchi, Danilo Alvares, Veronica Mericq, Ana Pereira, Susana Eyheramendy

## Abstract

Early or late pubertal onset can lead to disease in adulthood, including cancer, obesity, type 2 diabetes, metabolic disorders, bone fractures and psychopathologies. Thus, knowing the age at which puberty is attained is crucial as it can serve as a risk factor for future diseases. Pubertal development is divided into five stages of sexual maturation in boys and girls according to the standardized Tanner scale. We performed genome-wide association studies (GWAS) on the GOCS cohort composed of admixed children with European and Native American ancestry. Using joint models that integrate time-to-event survival parameters and longitudinal trajectories of body-mass index (BMI), we identified genetic variants associated with phenotypic transitions between pairs of Tanner stages. We identified 43 novel significant associations, most of them in boys. The GWAS on Tanner 3 *→* 4 transition in boys captured an association peak around the growth-related genes *LARS2* and *LIMD1* genes, the former of which causes ovarian dysfunction when mutated. The associated variants are expression– and splicing Quantitative Trait Loci regulating gene expression and alternative splicing in multiple tissues. Further, higher individual Native American genetic ancestry proportions predicted a significantly earlier arrival to Tanner 2 stage in boys but not in girls. Finally, the joint models identified longitudinal BMI parameters significantly associated in several Tanner stages’ transitions, confirming the association of BMI on pubertal timing.

## Introduction

Puberty is a complex process influenced by various factors such as genetics, nutrition, and environmental factors. Early or delayed onset of puberty can have significant consequences on long-term health outcomes and disease risk [1]. For example, early onset of puberty in girls has been linked to an increased risk of breast cancer, endometrial cancer, type 2 diabetes, cardiovascular diseases and psychopathologies later in life [1] [2] [3]. This is explained at least in part by a longer exposure to estrogen and insulin growth factor (IGF-1), which can affect cell proliferation and differentiation, as well as apoptosis [4]. Similarly, early puberty onset in boys has been associated with increased risk to obesity and high blood pressure in adulthood [1]. On the other hand, delayed puberty can lead to reduced adult height and decreased bone density, which can increase the risk of fractures and osteoporosis later in life [5] [6].

Pubertal maturation involves the appearance of a combination of physical changes that define secondary sexual characteristics in girls and boys. In girls, hallmark events are thelarche (the appearance of breasts), pubarche (the first appearance of pubic hair) and menarche (the first occurrence of menstruation), whereas in boys, hallmark events are changes in the scrotum and penis, pubarche, and increases in testicular volume. These sequences of events were described in seminal studies by Marshall and Tanner [7, 8], who defined 5 stages of maturation from prepubertal (Tanner stage 1) to fully mature (Tanner stage 5), with Tanner stage 2 representing the onset of puberty.

Pubertal maturation is influenced by diverse factors. One such factor is adiposity. Higher adiposity (i.e. BMI) correlates with an earlier age of onset of puberty, especially in girls [9] [10]. Indeed, BMI is synergistically related to pubertal timing as well as to other anthropometric traits like height [11, 12]. Thus, BMI is an important indicator of normal versus pathological sexual maturation [9, 13–16]. Ultimately, these processes are intertwined and regulated by shared endocrine mechanisms controlled by the hypothalamic-pituitary-gonadal system [17].

Ethnicity can also affect the variability of pubertal phenotypes [9]. For instance, the mean age of onset of breast development is higher in White American girls than in African American girls (10.0 and 8.9 years, respectively) [17]. Also, while menarcheal age is *>* 13 years old in northeastern European girls, in Latin American populations like Chile [18] and Venezuela is *∼* 12.5 years old. A study performed among Mapuche Indigenous and non-Indigenous girls from the Araucańıa region in Southern Chile showed that the age at menarche is 12.6 years in Mapuche girls, whereas it is 12.2 years in non-Indigenous girls (after correcting for socio-economic status) [19]. Further, Mapuche origin in Chile is an independent risk factor for precocious gonadarche (i.e. the earliest gonadal changes) and pubarche in boys but not in girls [20]. However, whether these differences are due to genetic ancestry or other factors is currently not well-understood [17].

Genetic factors explain 70-80% of the variance in pubertal timing, as revealed by monozygotic twin studies [17]. For instance, a large-scale GWAS on a European population identified 389 genetic variants associated with menarche, explaining *∼* 7.4% of the population variance [2]. In European boys, 76 variants were associated with the onset of facial hair (a marker of male puberty timing) [21].

The vast majority of genetic studies on pubertal variability have focused on cross-sectional phenotypes, namely, those that occur at specific time points (e.g. mean height at 8 years old), or the time point where a pubertal event takes place (e.g. age of menarche in girls). However, cross-sectional studies do not capture phenotypic changes over time. Instead, longitudinal models are suitable for analyzing phenotypic trajectories, such as BMI as a function of time. For instance, longitudinal GWAS have identified genetic variants associated with infant, child and/or adult BMI [22–24].

In this study, we investigated how genetic variability affects the transitions between sexual maturation stages. In particular, we were interested in estimating the length of time until the occurrence of a well-defined pubertal characteristic. We implemented survival models, which are best suited to analyse time-to-event observations. These models assume censored data as input [25–27]. There are different types of censorship, but we focused on interval and right censoring. The event is said to be interval censored when the event occurs within an interval of time, but the exact time of the event is unknown [28]. For example, in the context of this study, there was interval censorship in the data that records the stage number of the Tanner of a youth on each visit to the practitioner. A youth could be on Tanner 2 on one visit and in Tanner 3 on the following visit. Between the two visits, i.e. in that interval of time, he or she has changed from Tanner 2 to 3. On the other hand, the (very common) right censoring is when the event of interest is not observed in the last registration time or the patient has dropout of the study. This situation occurs, for example, when the youth in his/her last visit was not yet in Stage Tanner 5.

In many studies, longitudinal measurements are recorded along time-to-event data, but usually these two sources of data are analyzed separately. In certain settings, a joint modeling approach can benefit the analysis. For instance, when the association between longitudinal and survival outcomes is of interest [29] [30].

In this study, we implemented survival as well as joint models on data from admixed boys and girls with Native American and European ancestries. We were interested to assess whether some specific parameters from the BMI trajectories are associated with the transitions between Tanner stages. This can be studied in a joint model that combines the longitudinal modelling of BMI trajectories with time-to-event data fitting the transitions between Tanner stages.

In summary, the specific aims of this study are the following. First, to statistically model the transitions between Tanner stages. Second, to quantify how Native American genetic ancestry affects the timing of such transitions, when compared with European ancestry. Third, to identify genetic variants associated with the transitions. Forth, to assess the influence of BMI trajectories on the transition between Tanner Stages.

## Results

### Statistical modeling of transitions between Tanner stages

We studied the transitions between pairs of Tanner stages in boys and girls from the GOCS cohort. This cohort has longitudinal growth measurements from admixed Chilean children, collected for over 16 years [31]. We considered transitions from the consecutive Tanner stages pairs 1 *→* 2, Tanner 2 *→* 3 and Tanner 3 *→* 4. We also included the Tanner 2 *→* 4 transition because it encompasses the total duration of puberty, thus enabling us to analyse shared genetic factors across puberty (next sections). We implemented survival models, which estimate the probability S(t) at each age that an event of interest has not yet occurred. In our case the event of interest was the time point at which the Tanner stage changed, which occurred within an interval delimited by two consecutive visits to the practitioner. In the case of Tanner 1 *→* 2, the lower limit of the interval was set at the age of adiposity rebound (Age-AR; see Methods). **Fig 1** shows the survival curves obtained for Tanner 1 *→* 2, 2 *→* 3, 3 *→* 4 and 2 *→* 4 transitions in boys and girls. The parameter values of the model are shown in **S1 Table**. As expected, we observed that puberty in girls started and finished earlier than in boys (see **S2 Table** for the estimated ages at S(t) = 0.25, 0.5 and 0.75 in boys and girls). The progression from Tanner 2 *→* 3 was significantly slower in males than in females at most ages (**Fig 1** and **S3 Table**). For instance, at S(0.25), the Tanner 2 *→* 3 transition lasted 1.13 years in boys and 0.81 years in girls. **S3 Table** shows the time intervals between Tanner stages for S(t) = 0.25, 0.5 and 0.75.

**Fig 1.**
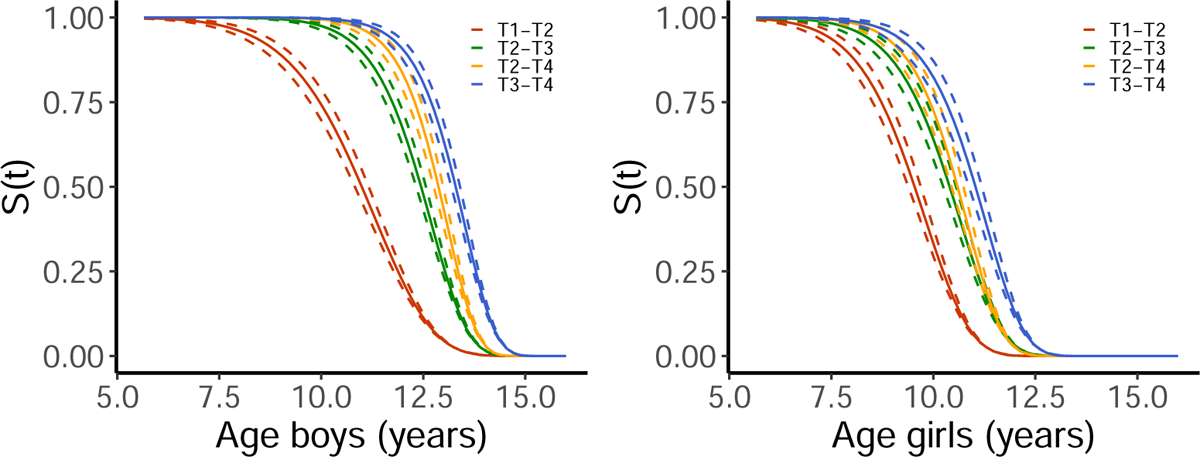
Survival curves for girls and boys. Probability S(t) of not moving to the next Tanner at a particular time for boys (left panel) and girls (right panel). Shown are the curves for Tanner 1 *→* 2, Tanner 2 *→* 3, Tanner 2 *→* 4, and Tanner 3 *→* 4 transitions.

### Effects of ancestry on transitions between Tanner stages

We estimated the ancestry proportions of the individuals from the admixed GOCS cohort (**S1 Fig**). We used the ADMIXTURE software, K=4 ancestral populations, and European, Native American and African reference populations (see Methods). These proportions were on average 52.1% European, 43.8% Mapuche Native American, 2.6% Aymara Native American, and 1.5% African. As explained above, we fitted a survival model to estimate the probability S(t) that at a given age a youth has not yet attained a particular Tanner stage given that he/she is in the previous Tanner stage. In this model, we adjusted for ancestry proportion. To highlight the effect of global ancestry, we considered survival curves from hypothetical individuals with 100% Mapuche ancestry and with 100% European ancestry, referred to as “Mapuche” and “Europeans” from now on. Global ancestry did have a significant effect in Tanner 1 *→* 2 transition in boys, with an effect size of 1.34 and a *P*-value = 0.026. However, global ancestry did not have a significant effect on the other transitions (**S1 Table**). **Fig 2** shows the survival curves of the Tanner 1 *→* 2 transition in Mapuche and European boys and girls. In quantitative terms, if we consider a 0.5 probability that boys have (not) attained Tanner 2 stage given that they are in Tanner 1, our model predicts that Mapuche and European boys will be 9.5 and 11.2 years old, respectively. **S4 Table** shows the predicted ages for probabilities S(t) of 0.25, 0.5 and 0.75, which follow a similar trend.

**Fig 2.**
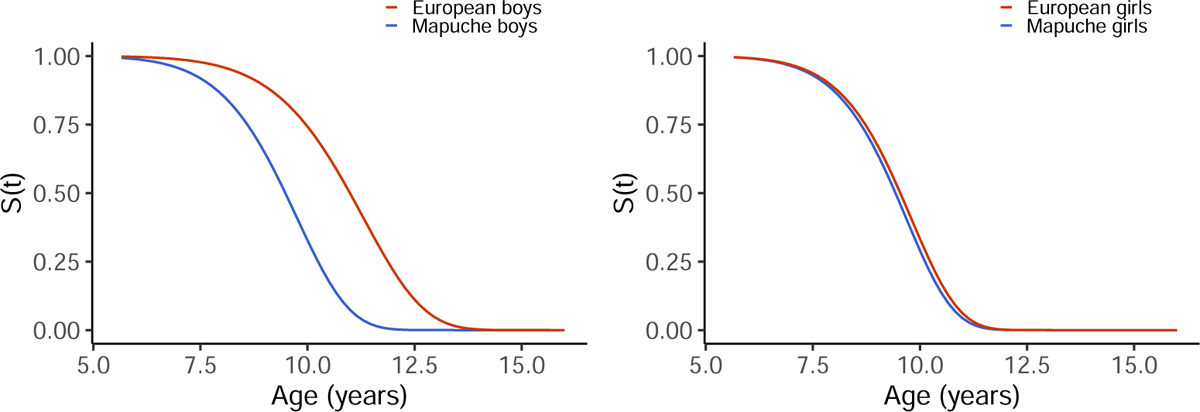
Effect of global ancestry on transitions between Tanner stages. Survival curves, which represent the probability S(t) of not moving to the next Tanner at a particular age. Shown are the curves for the Tanner 1 *→* 2 transition in Mapuche and European boys (left panel) and girls (right panel).

### GWAS on transitions between Tanner stages

We sought to identify single nucleotide polymorphism (SNP) alleles associated genome-wide with transitions between pairs of Tanner stages in boys and girls separately. We implemented survival models (see details in Methods) according to the following strategy. First, we ran GWAS with survival models for the interval censoring data consisting on Tanner stages between visits of the adolescents to the practitioner. In these models, the covariates used were the genetic variant, local ancestry for the SNP, and the global ancestry for the adolescent. Second, we implemented joint models that incorporate as a covariate a parameter from the longitudinal modeling of the log-BMI trajectories into the survival model (see details in Methods). The later step is crucial because, as mentioned before, BMI can affect the onset of puberty. Third, since running the GWAS based on the joint models on the whole array of SNPs is computationally expensive, we selected the SNPs from the survival analyses with association *P*-values lower than 0.1, obtaining between 49, 000 and 93, 000 SNPs (depending on the analysis).

The joint model’s captured 43 significant associations (*P*-value *<* 5 *×* 10*^−^*^8^) in total. **Table 1** shows these associations, together with the corresponding effect sizes and association *P*-values of the survival model, as well as functional annotations.

**Table 1.**
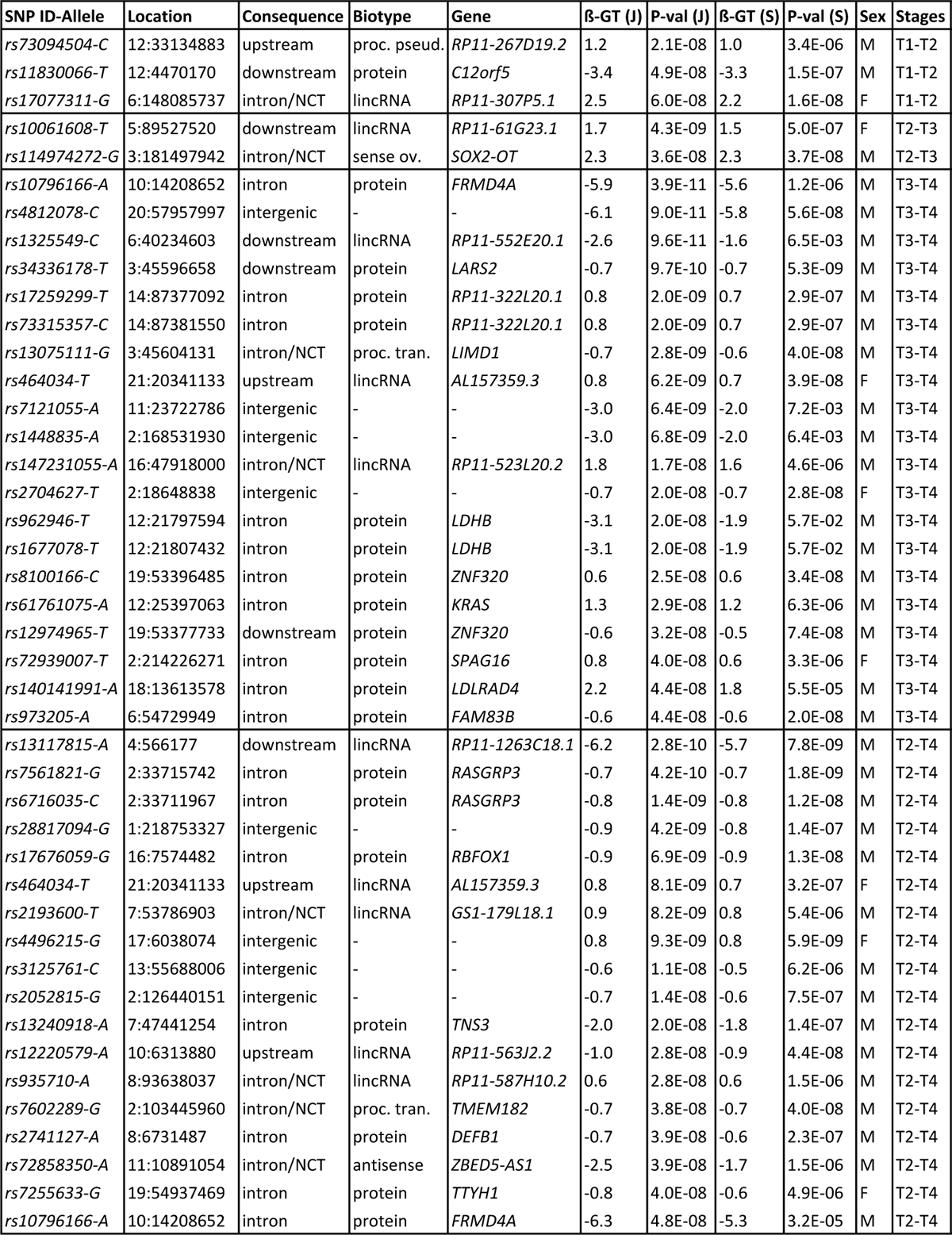
Significant GWAS associations for Tanner transition. Shown are the significantly associated variants identified by the joint models. SNP rs ID with the associated allele, physical location, sequence ontology (SO) consequence, biotype, gene, effect size of the genotype (*β_GT_*) in the joint (J) model, association *P*-value of the genotype (*P* _GT_) in the joint model, *β_GT_* in the survival (S) model, *P* _GT_ in the survival model, sex and Tanner transition stages. For each Tanner transition, variants were sorted in ascending order according to the *P* _GT_ in the joint model. M: male; F: female; NCT: noncoding transcript variant; proc.pseu.: processed pseudogene.

The GWAS for Tanner 1 *→* 2, Tanner 2 *→* 3, Tanner 3 *→* 4 and Tanner 2 *→* 4 transitions captured three, two, 20 and 18 significant associations, respectively. Out of them, 35 associations occurred in boys and 8 in girls. The 43 associations relate to 42 unique loci and 30 unique genes. The only SNP allele identified in two analyses was *rs464034-T*, which maps the long non-coding RNA *AL157359.3*, and was captured by the Tanner 2 *→* 4 and Tanner 3 *→* 4 GWAS in girls. No single SNP was significantly associated in both sexes in any analysis. Interestingly, the Tanner 3 *→* 4 GWAS in boys captured several variants forming peaks around genes *LARS2*, *LIMD1* (chromosome 3) and *FAM83B* (chromosome 6). The Manhattan plot in **Fig 3** shows the GWAS associations of the survival analysis, which display the association peaks. We do not show the Manhattans of the joint model’s GWAS because they were based on a reduced subset of SNPs. Significant associations surrounding less clear peaks also appear at chromosome 2 in boys and at chromosome 17 in girls in the Tanner 2 *→* 4 GWAS (**S4 Figure**). Most of the significantly associated SNPs from the remaining GWAS seem to drive the association alone (i.e. without linked variants). **S2 Figure**, **S3 Figure** and **S4 Figure** show the Manhattan plots for Tanner 1 *→* 2, Tanner 2 *→* 3 and Tanner 2 *→* 4 transitions from the survival models’ GWAS.

**Fig 3.**
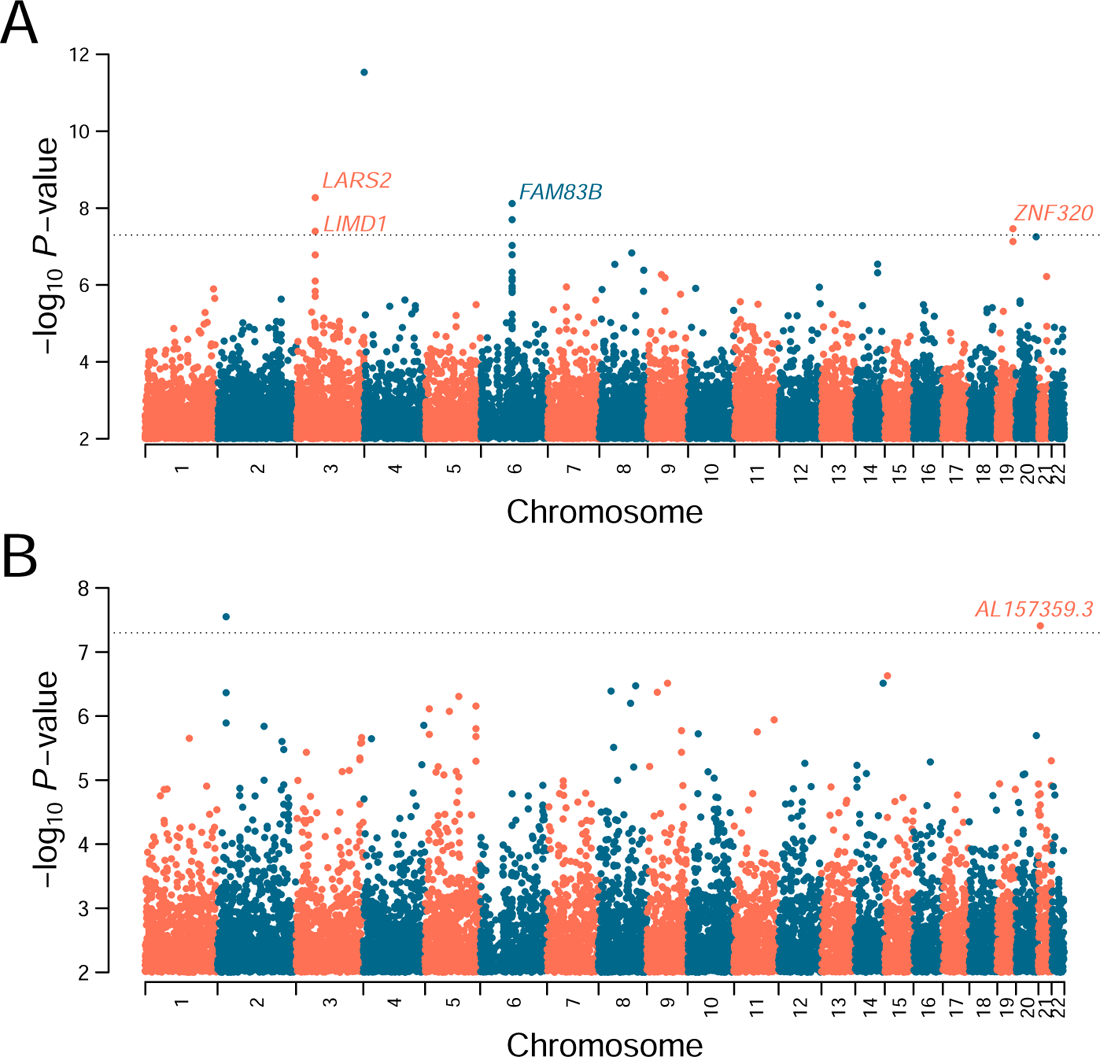
Survival GWAS on Tanner 3 *→* 4 transition. The Manhattan plots represent genotype-phenotype associations along the 22 autosomes in boys **(A)** and girls **(B)**. Highlighted are genes harboring variants whose associations were significant in both the joint and survival models’ GWAS. We show the plot made from the survival data, because the joint analysis was performed on a small subset of SNPs. The dotted black line represents the genome-wide significance threshold of *P* = 5 *×* 10*^−^*^8^. Variants with *−*log10(*P*-value) between 0 and 2 are not shown.

### Effect of longitudinal BMI on Tanner transitions

We assessed the effect of longitudinal BMI on Tanner transitions through a joint model (described in the Method Section). The joint model is a survival model that incorporates as a covariate a random effect from the modeling of the log-BMI trajectories through a mixed model. We assessed four different covariates to share from the random effects of the longitudinal mixed model. **S5 Table** shows the *P*-values for the association parameter when each of the four possible random effects were considered into the survival model. The four possible random effects considered were: the intercept, the slope, intercept plus the slope, and the mean (see details in the Methods Section). In all the joint models that we considered, the slope random effect was incorporated as a covariate. The effect of this covariate, called the association effect (or factor), was significant in boys in the Tanner 1 *→* 2 transition, with an effect size of 0.74 (*P* = 0.01) (**S1 Table**). In girls, the association factor was significant in Tanner 1 *→* 2 (effect size=0.9; *P* = 0.008), Tanner 3 *→* 4 (effect size=1.23; *P* = 0.002) and Tanner 2 *→* 4 (effect size=1.7; *P* = 1.6 *×* 10*^−^*^5^) (**S1 Table**).

### Functional annotations of associated genes

We queried the NHGRI GWAS Catalog [32] to identify reported GWAS associations for the genes detected in our analyses. We only considered reported genes with genome-wide association *P*-values lower than 5 *×* 10*^−^*^8^. Even though several genes have been previously GWAS-associated with diverse traits, we focused on those genes and traits more related to puberty. We defined three groups of trait categories: (i) Puberty, growth and endocrine function; (ii) obesity and anthropometric traits; and (iii) brain function (see Discussion). Each category was divided into several subcategories, and many genes belong to more than one (sub)category, reflecting pleiotropic effects. **Table 2** shows the identified genes, as well as their assigned categories and subcategories.

**Table 2.** Functional annotations of genes. Genes related to our GWAS were classified according to three trait categories and 29 subcategories related to puberty, based on published GWAS. According to existing variant and gene annotations, we classified the same genes into five categories. ^g^: gene associated to variant captured by our GWAS; ^e^: gene harboring eQTL captured by our GWAS; ^s^: gene harboring sQTL captured by our GWAS; ^E^: eGene of eQTL captured by our GWAS; ^S^: sGene of eQTL captured by our GWAS. Several genes were assigned to more than one category.

| Trait category | Trait subcategory | Genes |
| --- | --- | --- |
| Puberty, growth and endocrine function | Testosterone levels | *TNS3*<sup>g</sup>, *KRAS*<sup>g</sup>, *MFSD9*<sup>eE</sup>, *FGFRL1*<sup>s</sup> [35] |
|  | Sex hormone-binding globulin levels | *KRAS*<sup>g</sup> [36], *FGFRL1*<sup>s</sup> [35] |
|  | Age at first birth | *LARS2*<sup>gesES</sup> [37] |
|  | Age at onset of menarche | *FAM83B*<sup>ge</sup>, *RBFOX1*<sup>g</sup> [38], *WSCD1*<sup>eE</sup> [38] |
|  | Several male pubertal phenotypes | *SOX2-OT*<sup>g</sup> [21] [39] [40] [41] [42] |
|  | Adolescent idiopathic scoliosis | *SPAG16*<sup>ges</sup>, *TNS3*<sup>g</sup>, *TMEM182*<sup>g</sup>, *RBFOX1*<sup>g</sup>, *FRMD4A*<sup>g</sup> [43] |
|  | Heel bone mineral density | *FGFRL1*<sup>s</sup> [44] |
| Obesity and anthropometric traits | Type 2 diabetes | *LDLRAD4*<sup>g</sup> [45], *FRMD4A*<sup>g</sup> [46] |
|  | BMI | *LDLRAD4*<sup>g</sup>, *SOX2-OT*<sup>g</sup> [47], *LIMD1*<sup>gesES</sup> [48], *RBFOX1*<sup>g</sup> [49], *LARS2*<sup>gesES</sup> [50] |
|  | Waist-hip ratio | *ZBED5-AS1*<sup>gesE</sup> [38] |
|  | Cholesterol levels | *KRAS*<sup>g</sup> [51] |
|  | Body weight | *RBFOX1*<sup>g</sup>, *LDLRAD4*<sup>g</sup> [52] |
|  | Birth weight | *TNS3*<sup>g</sup> [53] |
|  | Height | *FRMD4A*<sup>g</sup>, *LIMD1*<sup>gesES</sup>, *LDHB*<sup>ge</sup> [38], *WSCD1*<sup>eE</sup> [54], *MFSD9*<sup>eE</sup>, *ZNF468*<sup>eE</sup>, *FGFRL1*<sup>s</sup> [55] |
|  | Body size or surface area | *LARS2*<sup>gesES</sup> [56], *FGFRL1*<sup>s</sup> [57] |
|  | Biological sex | *TMEM182*<sup>g</sup> [58] |
| Brain function | Educational attainment | *SPAG16*<sup>ges</sup>, *RBFOX1*<sup>g</sup> [59], *TMEM182*<sup>g</sup> [60], *TTYH1*<sup>gsS</sup> [61], *CDC42EP5*<sup>esE</sup> [60], *LENG8-AS1*<sup>eE</sup> [61], *LENG8*<sup>esES</sup> [60] |
|  | Cognitive ability | *SPAG16*<sup>ges</sup> [59], *RBFOX1*<sup>g</sup> [62] |
|  | Mathematical ability | *TMEM182*<sup>g</sup> [59] |
|  | Neuroticism | *RBFOX1*<sup>g</sup> [63] |
|  | Depression or bipolar disorder | *SPAG16*<sup>ges</sup> [64], *RBFOX1*<sup>g</sup> [65] |
|  | Anxiety | *RBFOX1*<sup>g</sup> [63] |
|  | Risk-taking behavior | *TMEM182*<sup>g</sup> [66] |
|  | Cannabis or cigarette smoking | *TMEM182*<sup>g</sup> [67] |
|  | Smoking initiation | *RBFOX1*<sup>g</sup> [68] |
|  | Decaffeinated coffee consumption | *RBFOX1*<sup>g</sup> [69] |
|  | Alcohol consumption | *FRMD4A*<sup>g</sup> [70], *WSCD1*<sup>eE</sup> [54] |
|  | Diet habits | *TMEM182*<sup>g</sup> [71] |
|  | Chronotype or morning person | *FAM83B*<sup>ge</sup> [72] |

An important proportion of disease heritability is explained by loci that regulate the expression of genes (“expression Quantitavite Trail Loci” or “eQTL”) and/or alternative splicing of pre-mRNA (“splicing Quantitative Trait Loci” or “sQTL”) [33]. Thus, we mined the Genotype-Tissue Expression (GTEx) [34] database to identify SNPs captured by our GWAS that are known eQTLs and/or sQTLs in single tissues. In total, we found 11 SNPs corresponding to eQTL and/or sQTL achieving significant associations in the Genotype-Tissue Expression (GTEx) project [34]. Among them, six are eQTLs only, one is a sQTLs only, and four are both eQTL and sQTL. The 11 eQTLs were significantly associated with the expression of 17 unique genes (“eGenes”) in at least one tissue (154 associations in total) (**S1 File**). The five sQTLs were significantly associated with the splicing of pre-mRNAs of 6 genes (“sGenes”; 83 associations in total) (**S2 File**). Next, we queried the NHGRI GWAS Catalog to identify previous puberty-related GWAS associations for the eGenes and sGenes. These genes, as well as genes harboring an eQTL or sQTL captured by our GWAS were assigned to the same categories and subcategories as before (**Table 2**).

## Discussion

In this study, we performed GWAS to identify genetic variants associated with the transitions between stages of pubertal maturation. Our methodological approach proved useful for capturing multiple genome-wide associations from a longitudinal cohort, even considering the small sample size of the GOCS cohort (**S6 Table**). To our knowledge, we are the first ones to implement this approach to dissect the genetic architecture of body growth, in particular of pubertal changes. The survival model’s GWAS alone detected more than 70 significant associations. However, after including a parameter from the BMI trajectories into the joint model, the total number of significant associations was reduced to 43 (see association *P*-values in **Table 1**). This indicates that some of the associations were mediated by BMI. Noteworthy, no gene or variant was shared between boys and girls, indicating a different genetic architecture between sexes. The only locus detected in more than one Tanner transition was the *AL157359.3* variant allele *rs464034-T*, which was captured by the Tanner 3 *→* 4 and Tanner 2 *→* 4 transitions in girls (**Table 1**). This suggests that this locus plays a role during the whole duration of female puberty, but that its effect sizes in Tanner 1 *→* 2 and Tanner 2 *→* 3 might be smaller than in the Tanner 3 *→* 4 transition. Future studies on a bigger cohort will be needed to test this hypothesis.

Among the 43 novel associations captured in our analyses, we identified genes and variants that had not been previously related to puberty. Perhaps the most relevant are *LARS2* and *LIMD1*. The associated variant alleles *rs34336178-T* and *rs13075111-G* top the peak at chromosome 3 in the Manhattan plot from the Tanner 3 *→* 4 transition in boys (**Fig 3**). *rs34336178-T* is located 1 bp downstream of *LARS2* and 228 bp of *LIMD1*. *LARS2* encondes the mitochondrial leucyl-tRNA synthetase, and enzyme that catalyzes the aminoacylation of tRNAs to bind their cognate amino acid during translation. Mutations in this gene cause Perrault syndrome, a rare autosomal recessive disorder which causes ovarian dysfunction in females, and bilateral sensorineural hearing loss in females and males [73]. *LIMD1* is a tumor-suppressor gene involved in a variety of functions, including gene expression regulation. Of note, *LARS2* and *LIMD1* have been GWAS-associated with obesity-related and anthropomorphic traits, such as BMI and height (**Table 2**). Importantly, *rs34336178* is a sQTL that regulates the alternative splicing of its own gene in six tissues, and it is also an eQTL that regulates its own expression, as well as *LIMD1*’s expression across 27 diverse tissues (**SFile1** and **SFile2**). The intronic/non-coding transcript variant *rs13075111* is an eQTL and sQTL regulating the expression of *LARS2*, *LIMD1* and/or *SACM1L* across 45 and 21 tissues, respectively (**SFile1** and **SFile2**). The observation that there are loci that work as eQTLs and sQTLs is expected, since alternative splicing can alter gene expression [33].

We also confirmed genes that had been GWAS-associated with pubertal phenotypes. One example is *SOX2-OT* (**S3 Figure**), which has been associated with pubertal and endocrine-related traits mostly in males, such as late vs. average onset facial hair, age at voice breaking [21], male-pattern baldness [39], excessive hairiness [40], monobrow [41] and eyebrow thickness [42]. The *SOX2-OT* variant allele *rs114974272-G* was detected in the Tanner 2 *→* 3 transition in boys. Interestingly, it is Tanner 3 stage where the spurt of pubic hair occur in boys [8]. *SOX2-OT* is a transcriptional regulator of *SOX2*, which is one of the major regulators of pluripotency [74]. *SOX2* is located within the sex-determining region of the Y chromosome (SRY) [75], which is necessary for male sex determination in mammals. Also, it is a key gene for the normal development and function of the hypothalamo-pituitary and reproductive axes, as revealed by clinical phenotypes of patients harboring heterozygous sequence variations in this gene [75]. We found three genes –*FAM83B*, *RBFOX1* and *WSCD1* – in boys that have been directly GWAS-associated with female puberty, in particular with the age at onset of menarche (**Table 2**). However, we did not find associations between these genes’ variants and Tanner transitions in girls. This result makes sense, since a proportion of menarche loci are important for pubertal initiation in boys as well [10]. The *FAM83B* intronic variant *rs973205*, which is part of the peak at chromosome 6 in the Manhattan plot of the Tanner 3 *→* 4 transition in boys (**Fig 3**), is an eQTL associated with the regulation of the undescribed *RP11-524H19.2* gene in the suprapubic skin, lower leg skin and esophageal mucosa (**SFile1** and **SFile2**).

Another interesting finding is the *TTYH1* intronic variant *rs7255633*, which was captured by the GWAS on Tanner 2 *→* 4 transition in girls. This variant is an eQTL and sQTL in 59 and 50 tissues, respectively. Its associated sGenes are *TTYH1*, *LENG8*, whereas its eGenes are *LENG8*, *CDC42EP5*, *LENG8-AS1*, *AC008746.12* and *CTD-2587H19.2*. Most of these genes have been GWAS-associated with educational attainment (**Table 2**), suggesting that *rs7255633* might play a role in brain functions related to sexual maturation across female puberty. The observation that many of the genes identified by our GWAS have known associations with brain function is in agreement with the critical bilateral effects between puberty and the brain. For example, our results show multiple associations with psychiatric conditions such as depression, bipolar disorder, anxiety, neuroticism or substance abuse (**Table 2**), and it is known that incidence rates of psychiatric disorders peak in adolescence [76].

We analysed and quantified how pubertal transitions are affected by Native American and European genetic ancestry. In all our survival and joint model we incorporated the global ancestry of each individual and local ancestry at each SNPs to adjust for population structure. Our results show that increased Mapuche proportions are significantly associated with an earlier arrival to Tanner 2 in boys. This result supports previous epidemiological findings in the GOCS cohort which show significant associations between Mapuche origin (based on the number of Mapuche last names) and precocious gonadarche and pubarche in boys [20]. While gonadarche refers to the earliest gonadal changes of puberty, pubarche refers to the first appearance of pubic hair at puberty. Moreover, a previous study by our group on the GOCS cohort found that the age at peak height velocity, namely, the age where the maximum rate of growth occurs during puberty (*∼* 12.7 years old among admixed Chileans), is 0.73 years earlier in Mapuche compared with European adolescents [77]. These observations provide further evidence that genetic ancestry is a relevant factor affecting the early physiological changes during puberty, particularly the first sexual maturation changes triggered in Tanner 2 stage.

A limitation of our study was that we could not replicate our findings in an independent cohort due to the lack of longitudinal pediatric growth cohorts with comprehensive data on Tanner stages, BMI trajectory and having a Native American ancestry component.

The results of this study are important to understanding the genetic architecture of pubertal changes, which hitherto has barely been analysed. In addition, they hold medical relevance, since they will serve to estimate with higher accuracy the genetic risk to potential associated diseases in adulthood. Finally, our results highlight the importance of implementing joint models of survival and longitudinal variables in GWA studies of growth processes, and hold great potential to get more insights into their complex genetic architecture.

## Methods

### Determination of Tanner stages

The “Growth and Obesity Chilean Cohort Study” (GOCS) [31] includes children with singleton births only, gestational age between 37 *−* 42 weeks, birth weight of 2, 500 to 4, 500 g, and no physical or psychological conditions that could severely affect growth. Among the 1196 participants of the cohort 943 (489 girls and 454 boys) met all inclusion criteria. To measure the sexual maturation of adolescents from GOCS, early clinical anthropometric evaluations were performed until 2009. Thereafter, at age 6.7 years, a single pediatric endocrinologist assessed breast, pubarche as well as genital development by palpation and classified breast, pubic hair and testes according to Tanner [7] [8]. Afterward, every 6 months, secondary sex characteristics were evaluated by a single dietitian trained for this purpose, with permanent supervision from the pediatric endocrinologist. Concordance between the dietitian and pediatric endocrinologist was 0.9 for breast and genitalia evaluation [31]. Age at menarche was self-reported. Since several boys did not wanted their testicules’ to be examined, Tanner 4 in males was labeled as the time of break voice. On the subset of boys that agreed to be examined, there was always an agreement between the ascertainment of Tanner 4 stage when labeled as break voice as well as testicular enlargement.

### Genotyping

We used SNP array data from GOCS adolescents obtained in a previous study [77]. Individuals were genotyped with the Infinium ^®^ Multhi-Ethnic Global BeadChip (Illumina). We used Plink 1.9 [78] to exclude 18 samples with call rate *<*0.98 (18 samples), 10 samples with gender mismatch, and one sample from each pair of highly related individuals (IBS *>*0.2). We excluded variants with a minor allele frequency (MAF) *<* 0.01, and variants following at least one of the following conditions: have heterozygous genotypes on male X chromosome, call genotypes on the Y chromosome in females, have high heterozigosity (*±* 3 SD from the mean), have *>*5% missing genotype data, have duplicated physical positions (one variant was kept from each duplicate pair) and show significant deviations from Hardy–Weinberg equilibrium (*P* = 1 *×* 10*^−^*^6^). We removed A-T and C-G transversions to avoid inconsistencies with the reference human genome. We also excluded 25 boys whose last BMI measurement was taken before they were 12 years old. After quality control filtering, we obtained the final data set of 904 individuals and 521, 788 autosomal SNPs.

### Local Ancestry Inference

We used RFmix.v.2.0 [79] to infer the local ancestry of each SNP allele from our Chilean sample. As reference populations we used Yoruba (YRI, n=108) for the African ancestry, Iberian Populations in Spain (IBS, n=107) for the European ancestry, Peruvian in Lima Peru (PEL) individuals with *>* 95% Native American ancestry (n=29). All of these samples are from the 1000 Genomes Project [80]. We excluded individuals with *>* 5% SNP missing rate. We estimated the haplotype phase from genotype data with Shapeit2 [81], using the HapMap37 human genome build 37 recombination map as a reference. We used the forward-backward parameter to run the software. RFmix identified three ancestries: Native American, European, and African.

### Global Ancestry Inference

Global ancestry proportions of Chilean children were estimated with Admixture 1.3.0 [82] in unsupervised mode. We used Yoruba (YRI, n=108) as the reference population for the African ancestry and Iberian Populations in Spain (IBS, n=107) for the European ancestry [80]. We used a merged dataset of 11 Mapuche [83] and 73 Aymara [84] [85] as the reference Native American population panel. Using K=4 the software clearly identified 4 ancestral groups corresponding to European, African, Aymara and Mapuche. The latter two groups represent the main Native American sub-ancestries of admixed Chileans. We used the Mapuche ancestry proportion as a covariate in the GWAS regressions. To distinguish Peruvian (PEL) individuals with *>* 95% Native American ancestry, we used K=3 ancestral groups and the aforementioned reference populations.

### Variant and gene annotations

SNP annotations were retrieved with the web server Variant Effect Predictor (VEP) from Ensembl [86], using the GRCh37 (hg19) human genome assembly. Upstream and downstream variants were defined as those located 10 Kb upstream or downstream of the gene, respectively. Intergenic variants were defined as those located *>* 100 Kb upstream or downstream of the closest gene. Reported GWAS associations were retrieved from the NHGRI GWAS Catalog [32]. We only considered genome-wide significant associations (*P <* 5 *×* 10*^−^*^8^). When more than one variant in the same gene has been associated with the same phenotype, we reported the strongest association. eQTLs and sQTLs data were retrieved using the Genotype-Tissue Expression (GTEx) Project [34] portal and variants achieving the significance threshold of *P* = 2.5 *×* 10*^−^*^7^ for eQTL mapping were considered significant [34].

### Derivation of the age of adiposity rebound

The derivation of Age-AR was described previously [87]. Before the derivation, 51 individuals were excluded because they had not enough measurements between 2 and 10 years old. Also, 156 individuals were excluded because all their measurements were taken after 4.5 years old. The final dataset had 696 individuals with measurements between 2 and 10 years old. The sample sizes for each analysis are displayed in **S6 Table**. To estimate Age-AR, the following longitudinal statistical model of BMI was implemented:

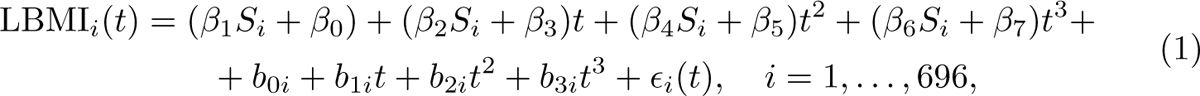

where LBMI*_i_*(*t*) represents the log-BMI of individual *i* at time/age *t* and *S_i_* represents the individual’s sex (0 female, 1 male). The parameters *β_k_* for *k* = 1, …, 7 are fixed effects whereas *b_ki_* for *k* = 0, …, 3 are random effects for each individual *i* where *b_ki_ ∼* Normal(0*, σ*^2^). The error terms *E_i_*(*t*) are assumed independent across the different individuals *i* but dependent between observations for the same individual and are normally distributed with mean at zero and variance *σ*^2^.

The predicted trajectory for individual *i* can then be written as

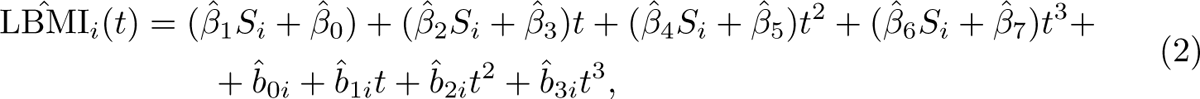

wherê(hat) is the estimator of the parameters. Age-AR is obtained by finding the time/age *t* at which the minimum BMI is found, by solving the equation:

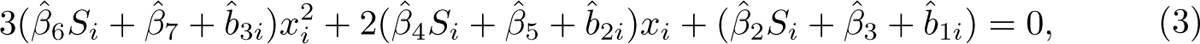

which has as its solution:

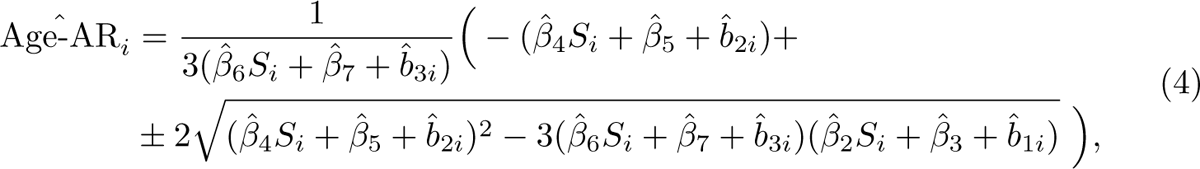

where the minimum obtained above needs to satisfy:

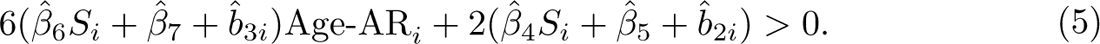

### GWAS for transitions between Tanner stages using a survival model

A survival model estimates the probability that a certain event has not occurred yet, which depends on some covariates. The hazard function represents the probability that the event occurred at time t.

Regardless of the type of censorship present in the data (interval or right censoring), the hazard function is usually estimated using a specification either parametric (e.g., Weibull baseline hazard) or semi-parametric (e.g., Cox proportional hazard) [88]. The hazard function for both approaches can be written, for an individual *i* at age *t*, as follows:

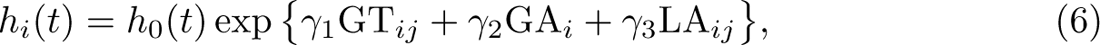

where *h*_0_(*t*) = *λ φ t^φ−^*^1^ specifies a Weibull baseline hazard while Cox’s proposal *h*_0_(*t*) is left unspecified. The following are the covariates in our models: GT*_ij_* is the additive representation of the genotype of SNP *j*, which takes values in *{*0, 1, 2*}*; GA*_i_* represents the global Mapuche Native American ancestry proportion; and LA*_i_* corresponds to the local ancestry that takes the values 0, 1 or 2 depending on the number of Native American alleles for SNP *j* and individual *i*. The effect of each variable is measured through its respective coefficient, ***γ*** = (*γ*_1_*, γ*_2_*, γ*_3_).

### Transitions between Tanner stages using a joint model of longitudinal and survival variables

Our joint modeling involves specifying two submodels, one longitudinal and one survival, that share information. The longitudinal model fits the trajectories of the logarithm of the BMI for each individual as a function of age using a mixed model. The survival model estimates the probability that individual *i* at age *t* has not yet attained Tanner stage *k*, for some *k ∈ {*2, 3, 4*}*, as a function of some covariates and some random effects estimated in the longitudinal models. By bringing random effects from the longitudinal model as covariates in the survival model we share information from the BMI trajectory of each individual. Different models can be specified by selecting a different set of random effects from the longitudinal model. After assessing several models we selected the model with the best performance according to the AIC criteria.

Specifically, the longitudinal submodel is a mixed model defined as follows [89]:

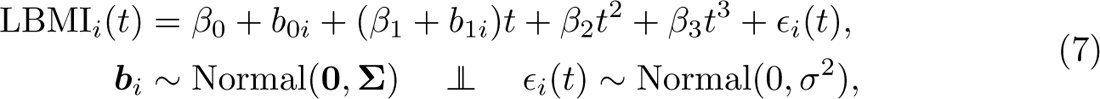

where LBMI*_i_*(*t*) represents the log-BMI of individual *i* at age *t*; (*β*_0_*, β*_1_*, β*_2_*, β*_3_) are intercept, linear, quadratic, and cubic fixed effects, respectively; ***b****_i_* = (*b*_0*i*_*, b*_1*i*_) are intercept and slope random effects, respectively; and *E_i_*(*t*) is an error term. **Σ** and *σ*^2^ are the variance-covariance matrix of random effects and the variance of error, respectively, and are parameters to be estimated.

The survival submodel (6) is defined as follows:

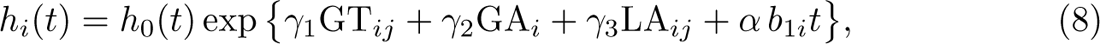

where GT*_ij_* is the additive representation of the genotype of SNP *j*at individual *i* taking values in *{*0, 1, 2*}*, GA*_i_* represents the global Mapuche Native American ancestry proportion, LA*_ij_* corresponds to the local ancestry and takes the values 0, 1 or 2 depending on the number of Native American alleles for SNP *j* and individual *i*. *α* quantifies the strength of the association between the longitudinal and survival processes, the so-called association effect. Note that we are only sharing the time-dependent random effect (i.e., the slope). In our context, we are interested in the association of this effect (BMI trend) with time to puberty.

Estimating the parameters and random effects simultaneously is a computationally demanding task. In particular, marginal likelihood is required (i.e., integrating out the random effects), making the inference even more expensive and impractical for large datasets. For these reasons, we implement the procedure proposed by [90], which consists of two-stage inference: (1) the longitudinal submodel is fitted, and (2) the estimation of *b*_1*i*_ is used as a covariate into the survival submodel.

## Consent to Participate

Informed consent from participants was obtained from parents or guardians. Children agreed to participate when they turned 7 years old. This study was approved by the Scientific Ethics Committees of Instituto de Nutricíon y Tecnoloǵıa en Alimentos (INTA) and Pontificia Universidad Católica de Chile.

## Author Contributions

S.E. conceived the study. S.E., L.V. and D.A. designed experiments. E.B., L.V. and V.L-Y. performed analyses. V.M. and A.P. performed anthropometric measurements and collected medical data. L.V., S.E. and D.A. wrote the manuscript. All the authors critically reviewed and accepted the final version.

## Data Availability Statement

The genetic data used in this study are from adolescents, many of which are less than 18 years old. Thus, we are not allowed to publish or share their raw data. The summary statistics of this study can be found in the NHGRI-EBI GWAS Catalog server [32], under the code GCP000453.

## Data Availability

The genetic data used in this study are from adolescents, many of which are less than 18 years old. Thus, we are not allowed to publish or share their raw data. The summary statistics of this study can be found in the NHGRI-EBI GWAS Catalog server, under the code GCP000453.

## Supplementary Material

### Supplementary Figures

**S1 Figure.**
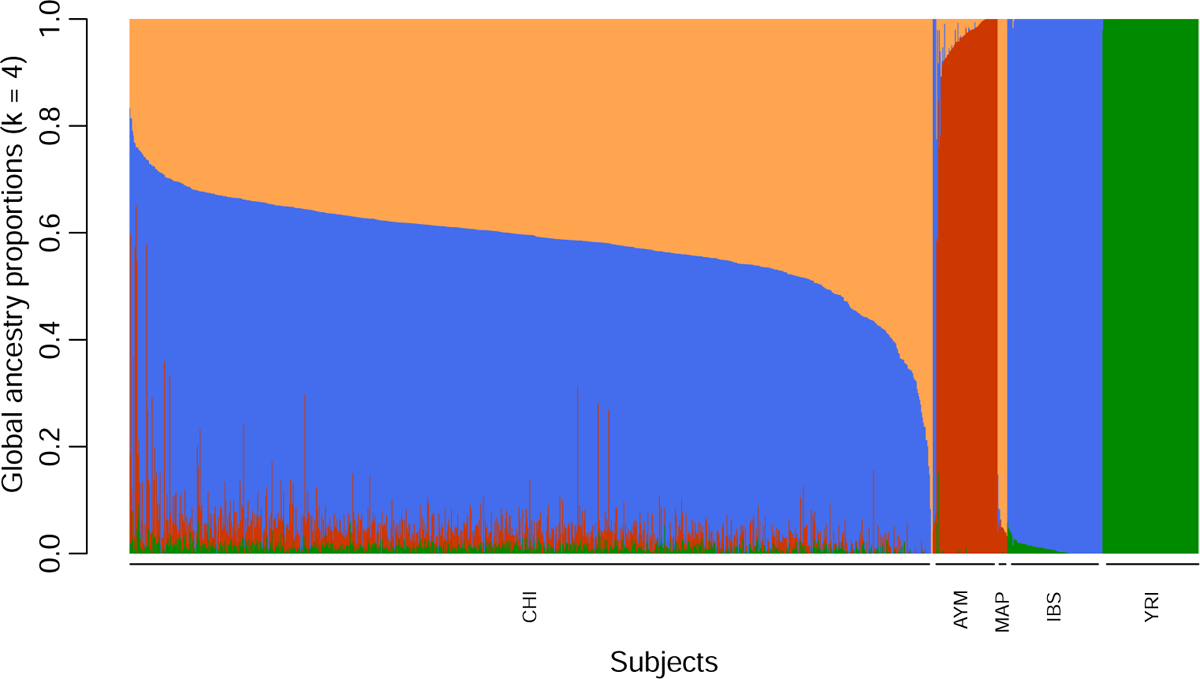
Global ancestry proportions of admixed Chilean individuals. The number of ancestral populations used was *K* = 4. CHI: Admixed Chileans; AYM: Aymara; MAP: Mapuche; IBS: Spaniards; YRI: Yoruba.

**S2 Figure.**
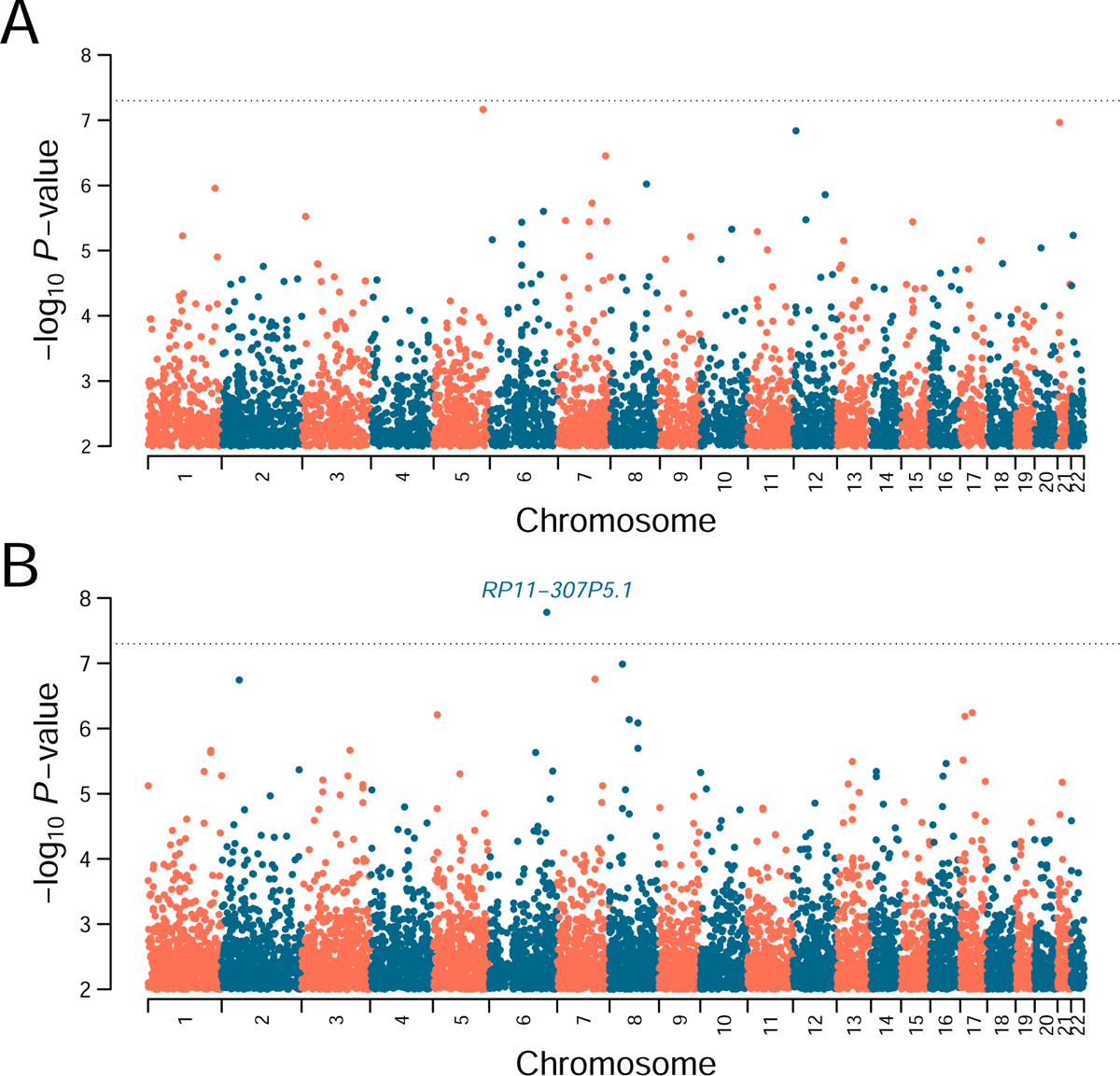
GWAS of Tanner 1 *→* 2 transition captured by the survival model. The Manhattan plots represent genotype-phenotype associations along the 22 autosomes in boys **(A)** and girls **(B)**. Highlighted are genes associated with variants whose associations were significant in the joint and survival models’ GWAS. The dotted black line represents the genome-wide significance threshold of *P* = 5 *×* 10*^−^*^8^. Variants with *−*log10(*P*-value) between 0 and 2 are not shown.

**S3 Figure.**
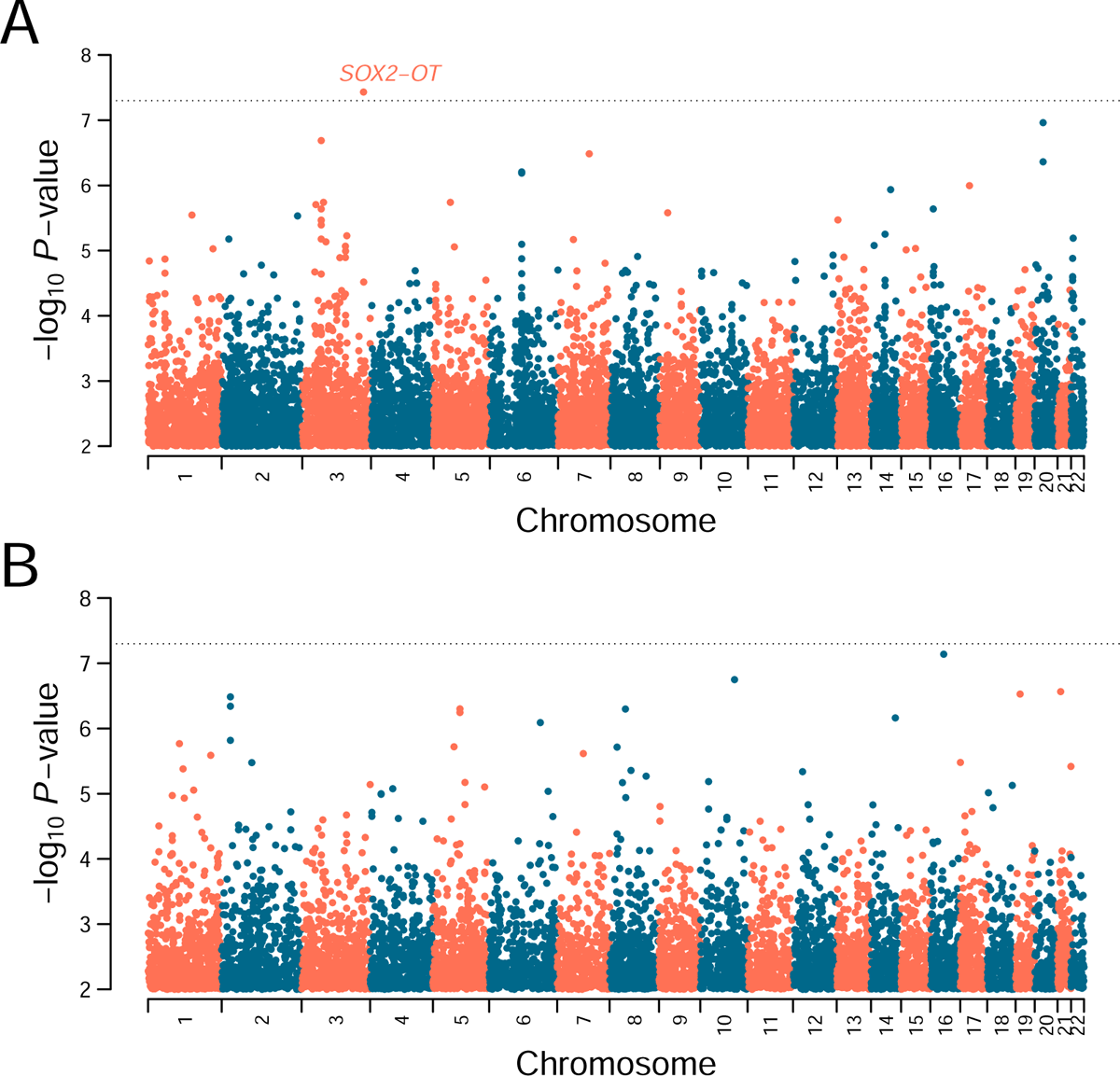
GWAS of Tanner 2 *→* 3 transition captured by the survival model. The Manhattan plots represent genotype-phenotype associations along the 22 autosomes in boys **(A)** and girls **(B)**. Highlighted are genes associated with variants whose associations were significant in the joint and survival models’ GWAS. The dotted black line represents the genome-wide significance threshold of *P* = 5 *×* 10*^−^*^8^. Variants with *−*log10(*P*-value) between 0 and 2 are not shown.

**S4 Figure.**
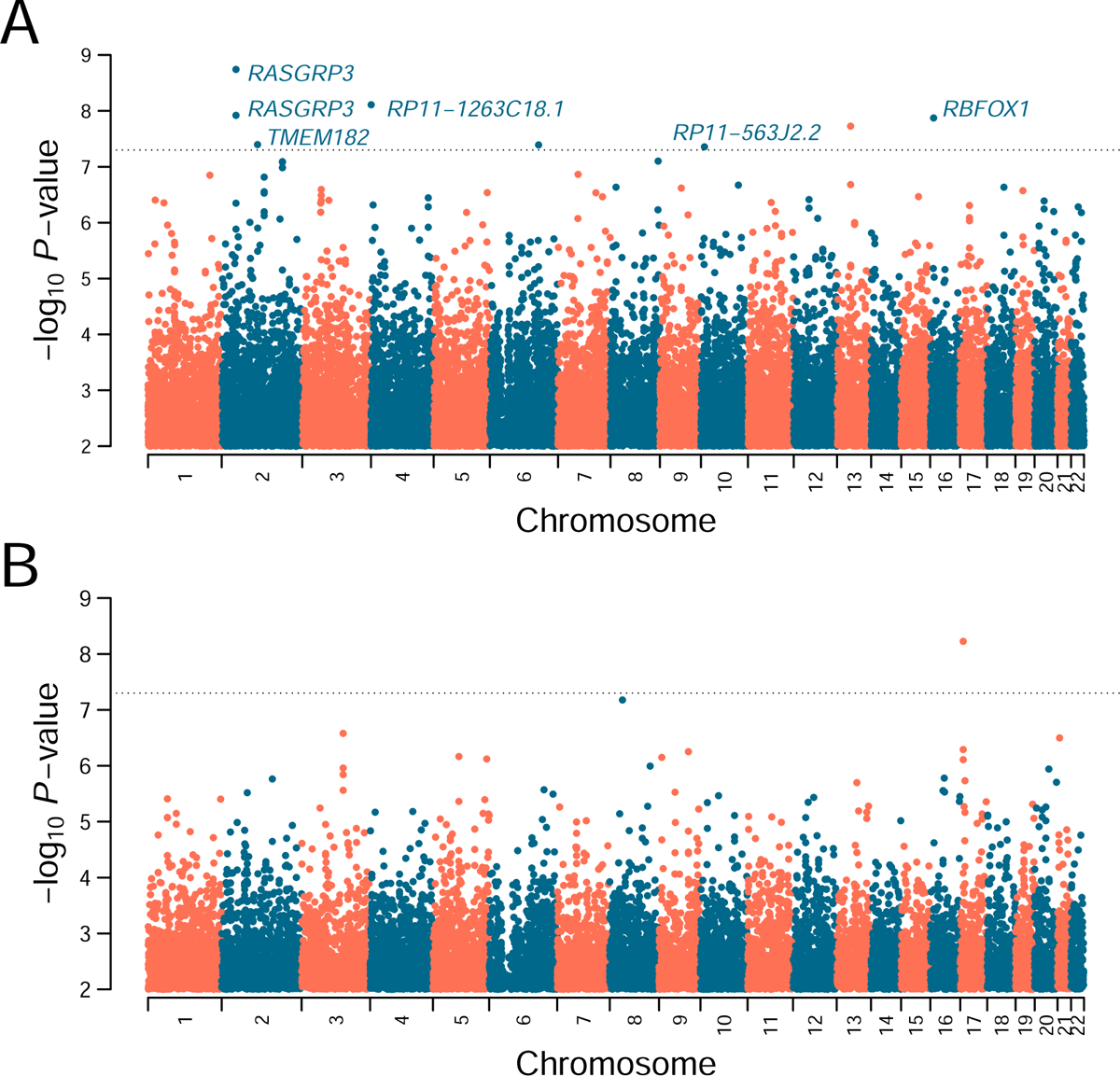
GWAS of Tanner 2 *→* 4 transition captured by the survival model. The Manhattan plots represent genotype-phenotype associations along the 22 autosomes in boys **(A)** and girls **(B)**. Highlighted are genes associated with variants whose associations were significant in the joint and survival models’ GWAS. The dotted black line represents the genome-wide significance threshold of *P* = 5 *×* 10*^−^*^8^. Variants with *−*log10(*P*-value) between 0 and 2 are not shown.

### Supplementary Tables

**S1 Table.** Summary statistics for global ancestry and BMI association of the joint model by Tanner transition. Shown are the variables and their respective effect size, standard error (SE), and *P*-value at each Tanner stage transition according to sex.

| Sex | Tanner Trans. | Variable | Effect Size | SE | P-value |
| --- | --- | --- | --- | --- | --- |
| Girls | $T_1 \rightarrow T_2$ | Global ancestry | -0.120 | 0.734 | 0.870 |
|  |  | Assoc. Factor | 0.901 | 0.341 | 0.008 |
| | $T_2 \rightarrow T_3$ | Global ancestry | -0.199 | 0.852 | 0.815 |
|  |  | Assoc. Factor | 0.587 | 0.311 | 0.059 |
| | $T_3 \rightarrow T_4$ | Global ancestry | 0.221 | 0.814 | 0.786 |
|  |  | Assoc. Factor | 1.233 | 0.401 | 0.002 |
| | $T_2 \rightarrow T_4$ | Global ancestry | 0.166 | 0.851 | 0.845 |
|  |  | Assoc. Factor | 1.709 | 0.393 | <0.001 |
| Boys | $T_1 \rightarrow T_2$ | Global ancestry | 1.321 | 0.621 | 0.033 |
|  |  | Assoc. Factor | 0.739 | 0.287 | 0.010 |
| | $T_2 \rightarrow T_3$ | Global ancestry | 0.942 | 0.61 | 0.128 |
|  |  | Assoc. Factor | 0.429 | 0.317 | 0.176 |
| | $T_3 \rightarrow T_4$ | Global ancestry | 0.732 | 0.639 | 0.252 |
|  |  | Assoc. Factor | 0.496 | 0.314 | 0.114 |
| | $T_2 \rightarrow T_4$ | Global ancestry | 0.337 | 0.786 | 0.668 |
|  |  | Assoc. Factor | 0.411 | 0.407 | 0.313 |

**S2 Table.** Mean ages attained at Tanner transitions. Shown are the Tanner pair, the value (probability) of the survival function S(t), the median age attained at the Tanner transitions in males (M) and females (F), including the confidence intervals’ lower (L) and upper bounds (U).

| Tanner Trans. | S(t) | Age M (L) | Age M | Age M (U) | Age F (L) | Age F | Age F (U) |
| --- | --- | --- | --- | --- | --- | --- | --- |
| $T_1 \rightarrow T_2$ | 0.25 | 11.7 | 12.0 | 12.2 | 10.1 | 10.3 | 10.6 |
| $T_2 \rightarrow T_3$ | 0.25 | 12.9 | 13.1 | 13.3 | 10.9 | 11.2 | 11.3 |
| $T_2 \rightarrow T_4$ | 0.25 | 13.2 | 13.4 | 13.6 | 11.1 | 11.3 | 11.5 |
| $T_3 \rightarrow T_4$ | 0.25 | 13.7 | 13.9 | 14.1 | 11.6 | 11.8 | 12.0 |
| $T_1 \rightarrow T_2$ | 0.50 | 10.7 | 11.2 | 11.6 | 9.2 | 9.7 | 10.2 |
| $T_2 \rightarrow T_3$ | 0.50 | 12.2 | 12.6 | 13.1 | 10.0 | 10.5 | 11.0 |
| $T_2 \rightarrow T_4$ | 0.50 | 12.4 | 12.9 | 13.4 | 10.2 | 10.7 | 11.1 |
| $T_3 \rightarrow T_4$ | 0.50 | 12.9 | 13.4 | 13.9 | 10.7 | 11.2 | 11.6 |
| $T_1 \rightarrow T_2$ | 0.75 | 9.3 | 10.0 | 10.7 | 8.0 | 8.7 | 9.4 |
| $T_2 \rightarrow T_3$ | 0.75 | 11.1 | 11.8 | 12.5 | 9.0 | 9.7 | 10.4 |
| $T_2 \rightarrow T_4$ | 0.75 | 11.7 | 12.4 | 13.1 | 9.3 | 10.0 | 10.8 |
| $T_3 \rightarrow T_4$ | 0.75 | 12.0 | 12.8 | 13.5 | 9.6 | 10.3 | 11.0 |

**S3 Table.** Time intervals between survival curves. Shown are pairs of Tanner stages’ pairs representing the curves, the value (probability) of the survival function S(t) and the time interval between the curves in males and females. * Significant association.

| Interval | S(t) | Males (years) | Females (years) |
| --- | --- | --- | --- |
| T12 - T23 | 0.25 | 1.13* | 0.81* |
| T12 - T23 | 0.50 | 1.45* | 0.81 |
| T12 - T23 | 0.75 | 1.78* | 0.97 |
| T23 - T34 | 0.25 | 0.81* | 0.65* |
| T23 - T34 | 0.50 | 0.81 | 0.65 |
| T23 - T34 | 0.75 | 0.97 | 0.65 |
| T23 - T24 | 0.25 | 0.32* | 0.16 |
| T23 - T24 | 0.50 | 0.32 | 0.16 |
| T23 - T24 | 0.75 | 0.65 | 0.32 |
| T24 - T34 | 0.25 | 0.48* | 0.48* |
| T24 - T34 | 0.50 | 0.48 | 0.48 |
| T24 - T34 | 0.75 | 0.32 | 0.32 |

**S4 Table.** Age attained at Tanner *T*_1_ *→ T*_2_ **transition in Mapuche and European boys.** Shown are the median age and its confidence intervals’ lower (L) and upper (U) bounds for S(t) = 0.25, 0.5 and 0.75.

| Ancestry | S(t) | Age (L) | Age (median) | Age (U) |
| --- | --- | --- | --- | --- |
| Mapuche | 0.25 | 9.37 | 10.34 | 11.64 |
|  | 0.5 | 8.73 | 9.54 | 10.83 |
|  | 0.75 | 7.92 | 8.73 | 9.70 |
| European | 0.25 | 11.80 | 11.96 | 12.12 |
|  | 0.5 | 10.83 | 11.15 | 11.31 |
|  | 0.75 | 9.86 | 10.02 | 10.34 |

**S5 Table.** *P*-value for association parameter of the joint model by Tanner transition. Shown are the P-values for association parameters of the joint model with different shared component specifications at each Tanner stage transition according to sex.

| Sex | Tanner Trans. | Intercept | Slope | Intercept + Slope | Mean |
| --- | --- | --- | --- | --- | --- |
| Girls | $T_1 \rightarrow T_2$ | 0.881 | 0.008 | 0.004 | 0.005 |
| | $T_2 \rightarrow T_3$ | 0.828 | 0.057 | 0.020 | 0.025 |
| | $T_3 \rightarrow T_4$ | 0.735 | 0.001 | 0.001 | 0.002 |
| | $T_2 \rightarrow T_4$ | 0.333 | <0.001 | <0.001 | <0.001 |
| Boys | $T_1 \rightarrow T_2$ | 0.478 | 0.007 | 0.016 | 0.020 |
| | $T_2 \rightarrow T_3$ | 0.371 | 0.015 | 0.063 | 0.062 |
| | $T_3 \rightarrow T_4$ | 0.598 | 0.089 | 0.066 | 0.056 |
| | $T_2 \rightarrow T_4$ | 0.488 | 0.284 | 0.558 | 0.511 |

**S6 Table.** Sample sizes of survival and joint analyses. Shown are the sample sizes for girls and boys.

| Sex | Tanner Transition | N Survival | N Joint |
| --- | --- | --- | --- |
| **Girls** | $T_1 \rightarrow T_2$ | 332 | 292 |
| | $T_2 \rightarrow T_3$ | 254 | 247 |
| | $T_3 \rightarrow T_4$ | 212 | 211 |
| | $T_2 \rightarrow T_4$ | 252 | 251 |
| **Boys** | $T_1 \rightarrow T_2$ | 383 | 381 |
| | $T_2 \rightarrow T_3$ | 346 | 346 |
| | $T_3 \rightarrow T_4$ | 323 | 323 |
| | $T_2 \rightarrow T_4$ | 318 | 318 |

### Supplementary Files’ Legends

**SFile1. eQTLs associated with the** 42 **loci captured by our GWAS.** Shown are the SNP rs ID, physical location, association P-value, effect size, gene ID and eGene ID.

**SFile2. sQTLs associated with the** 42 **loci captured by our GWAS.** Shown are the SNP rs ID, physical location, association P-value, effect size, gene ID and sGene ID.

